# Genome-wide tandem repeat expansions contribute to schizophrenia risk

**DOI:** 10.1101/2021.12.17.21267642

**Authors:** Bahareh A Mojarad, Worrawat Engchuan, Brett Trost, Ian Backstrom, Yue Yin, Bhooma Thiruvahindrapuram, Linda Pallotto, Mahreen Khan, Giovanna Pellecchia, Bushra Haque, Keyi Guo, Tracy Heung, Gregory Costain, Stephen W Scherer, Christian R Marshall, Christopher E Pearson, Anne S Bassett, Ryan KC Yuen

## Abstract

Tandem repeat expansions (TREs) can cause neurological diseases but their impact in schizophrenia is unclear. Here we analyzed genome sequences of adults with schizophrenia and found that they have a higher burden of TREs that are near exons and rare in the general population, compared with non-psychiatric controls. These TREs are disproportionately found at loci known to be associated with schizophrenia from genome-wide association studies, in individuals with clinically-relevant genetic variants at other schizophrenia loci, and in families where multiple individuals have schizophrenia. Our findings support the involvement of genome-wide rare TREs in the polygenic nature of schizophrenia.

## Main text

Schizophrenia is a major neuropsychiatric disorder with heritability estimated at ∼79%^1^. Previous studies have revealed the role of copy number variants (CNVs)^2,3^ and small nucleotide variants in its pathogenesis^4^. However, those analyses were not designed to interrogate repetitive regions of the genome owing to challenges in detecting and interpreting such variation. In a separate study^5^, we used a comprehensive analytic strategy to investigate tandem DNA repeats, which constitute ∼6% of the human genome, and showed that genome-wide TREs are associated with the risk of autism spectrum disorder (ASD), a complex neurodevelopmental disorder with genetic risk that overlaps that of schizophrenia^6^. Our recent genome sequence analysis provided evidence supporting the involvement of TREs in schizophrenia through the identification of potentially damaging TREs in known disease-associated loci from a cohort of unrelated adults with schizophrenia^7^.

Here, we used ExpansionHunter Denovo (EHdn)^8^ and our established analytic approach to analyze TREs in the genomes of 257 unrelated adult cases with schizophrenia of European ancestry, 225 ancestry- and sequence-pipeline-matched individuals with no major neuropsychiatric disorders (non-psychiatric controls)^7^, and in 2,504 individuals from the 1000 Genomes Project^9^ to estimate population frequency of TREs in cases and controls (**Methods**). Our approach interrogates the entire genome irrespective of prior knowledge of the presence or expected sequence of tandem repeats in any given region, and focuses on tandem repeats having motifs of 2-20 bp for which the total repeat tract length is greater than the sequencing read length (i.e., >150 bp)^8^. We define a tandem repeat to be expanded when its tract length is an outlier compared to lengths at that loci in other individuals^5^ (**Methods**).

After quality assessment and parameter optimization (**Methods**), we performed a burden analysis comparing rare TREs (<0.5% frequency in 1000 Genomes Project individuals) in individuals with schizophrenia and in non-psychiatric controls. We identified 583 rare TREs in 436 distinct regions in 220 individuals with schizophrenia (**Supplementary Table S1**); 199 of these distinct regions were genic (involving 193 genes, hereafter referred to as TRE-associated genes, including 6 genes with multiple repeat motifs/regions identified). In individuals with schizophrenia, rare TREs tend to be located within genes (**Figure 1A**), and more likely to be at exon junctions (OR=5.03, p=6.0×10^−3^, **Figure 1B**). Fine mapping of the eight exon-proximal TREs revealed their precise locations to be in intronic or untranslated regions with close proximity (<300 bp) to protein-coding exons. This included a CTG expansion in myotonic dystrophy-linked *DMPK* we reported previously^7^ (**Supplementary Information** and **Supplementary Table S2**).

**Figure 1.**
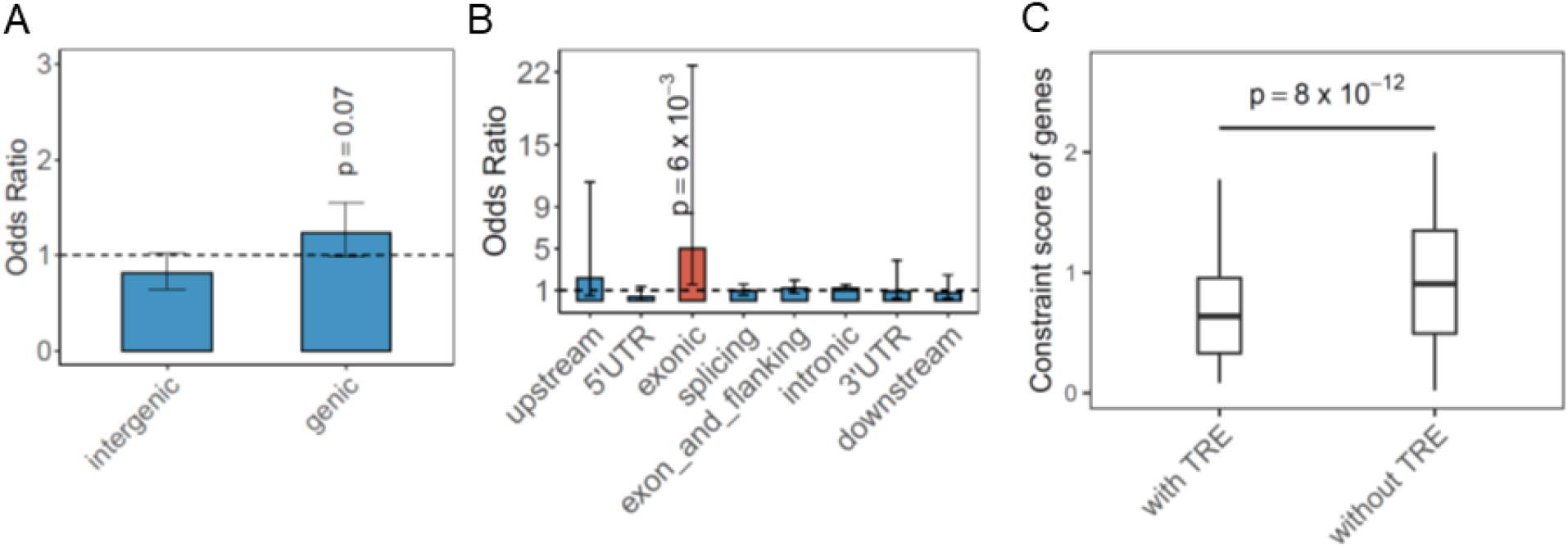
Genetic features and functional impact of rare TREs. **A)** Burden analysis of rare TREs located near (genic) or outside (intergenic) genes. **B)** Burden analysis of rare TREs with different genomic features in individuals with schizophrenia relative to non-psychiatric controls. Red bar indicates significant enrichment of exon-proximal (labelled as exonic) rare TREs in individuals with schizophrenia. Horizontal dashed line represents odds ratio=1. **C)** Distributions of gnomAD observed/expected (o/e) upper bound LOEUF values are shown for 182 genes with rare TREs (182 of 193 TRE-associated genes have scores) in the 220 (of 257) individuals with schizophrenia, compared with 18,990 genes with no such TREs identified in the schizophrenia cohort (one-sided Wilcoxon rank-sum test). Minima and maxima indicate 3× the interquartile range-deviated o/e upper bounds from the median and the centre indicates the median of the o/e upper bound values.

In 139 of the individuals with schizophrenia, there were 222 rare intronic TREs in 160 distinct regions of 155 genes (including 5 genes with multiple repeat motifs/loci), representing the largest subcategory of the genic region. Of these, 51 individuals had rare intronic TREs in one or more of the 38 genes that are associated with neurological abnormality, abnormal behavior or nervous system abnormality in Mammalian Phenotype Ontology, including genes previously associated with schizophrenia such as *DCLK1, ERBB4, GRIK4, GRIN2A, SHANK1*, and *VIPR2* (**Supplementary Table S3**). Gene-set analysis of rare exon-proximal and intronic TREs identified a significant enrichment of genes involved in postnatal brain expression, such as *GRIN2A* and *SHANK1* (OR=1.83, p=8.6×10^−3^, false discovery rate=0.2) (**Supplementary Table S4** and **Supplementary Figure 1**). TRE-associated genes were significantly more constrained than other genes as measured by the GnomAD loss-of-function observed/expected upper bound fraction (LOEUF)^10^ (p=3×10^−10^) (**Figure 1C**), suggesting that these schizophrenia-associated rare TREs may impact genes in a loss-of-function manner.

Our previous analysis of this same community-based cohort of adults with schizophrenia included 33 individuals with clinically relevant CNVs and small nucleotide variants, constituting 12.8% of individuals studied^7,11,12^. Of the 13 individuals with the eight rare exon-proximal TREs, five had at least one other clinically relevant (non-TRE) rare variant (i.e., small nucleotide/copy number variants) (**Supplementary Table S5**)^7,11,12^; a significant association (p=3.56×10^−3^) (**Figure 2A, and Supplementary Information**). This supports schizophrenia as a complex disorder involving multiple genetic risk factors^13^.

**Figure 2.**
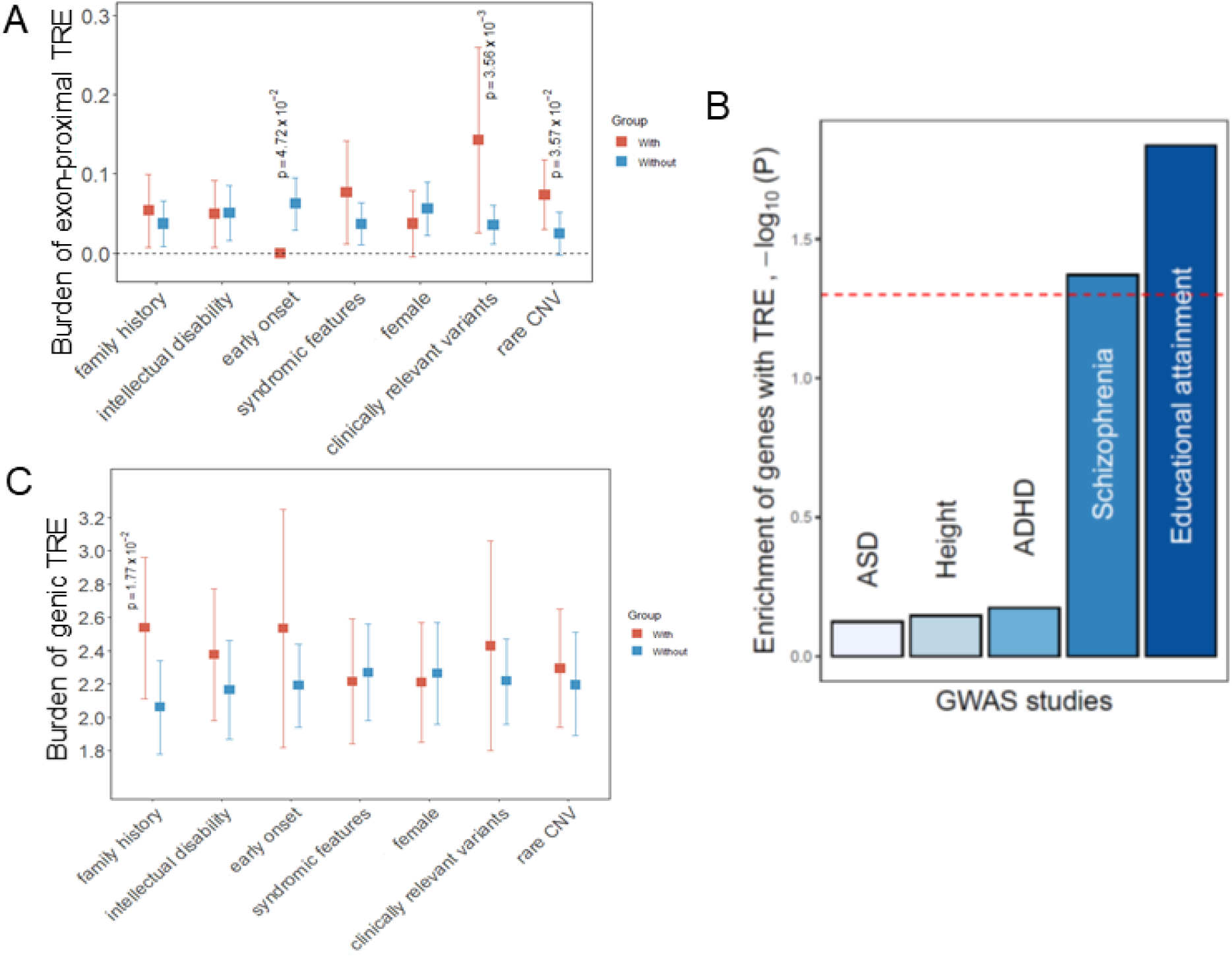
Genetic and clinical features involved in rare genic TREs in schizophrenia. The burden of **A)** rare exon-proximal and **C)** rare genic TREs in schizophrenia was analyzed with respect to presence/absence of seven variables (x-axis). Orange and blue coloured boxes indicate results for TRE-containing individuals, with and without each of the seven variables, respectively, with vertical bars representing 95% confidence intervals; p-values are provided above variables noting significant differences. No individuals with a rare exon-proximal TRE had an early age at onset of schizophrenia. **B)** MAGMA was used to assess the 193 genes with rare TREs detected by our pipeline against proximity (<10 kb) to common risk variants from GWAS studies for schizophrenia, ASD, attention deficit/hyperactivity disorder, educational attainment, and (as a negative control) height (**Methods**). The dashed red line represents association with p-value equal to 0.05.

Next, we used MAGMA^14^ to integrate summary statistics from genome-wide association studies (GWASs) of five traits (ASD, height, attention deficit hyperactivity disorder, schizophrenia and educational attainment), and examined whether common genetic variation influencing thess traits were located within 10 kb of the 193 TRE-associated genes we identified (**Methods**). We determined that signals for schizophrenia and for educational attainment (where the GWAS signal also correlates with schizophrenia), but not the other three GWAS signals tested (**Methods**), showed significant enrichment for our TRE-associated genes (**Figure 2B**), further supporting their contribution to the polygenic risk of schizophrenia. Eleven of the 193 TRE-associated genes, including *DPYD* and *EFNA5*, were disproportionally found amongst the 655 genes potentially tagged by genome-wide significant common variant signals at 270 loci in the latest schizophrenia GWAS (OR=2.65, p=5×10^−3^), but only two (*GRIN2A* and *MYT1L*) were in the 114 protein-coding genes prioritized using current variant mapping and expression methods.^15^

In our schizophrenia cohort, we also found that individuals with a family history of schizophrenia were more likely to carry rare TREs in genic regions (p=1.77×10^−2^) (**Figure 2C**), suggesting that these individuals may have inherited the rare TREs. To investigate the involvement of rare genic TREs in families with a history of schizophrenia, we performed genome sequencing on 63 additional individuals (30 of whom were diagnosed with schizophrenia) from 14 independent families with an extended family history of schizophrenia^16^. In two probands of these 14 families, we detected rare TREs in three genic regions (*CALCOCO2* and *FXN* in one family, and *SHANK1* in the other family) that were also identified in our primary schizophrenia cohort (**Supplementary Table S6**). In these two families, all five genome-sequenced individuals with schizophrenia carried the detected rare genic TREs, even though the same expansions can be found in some of the unaffected family members (**Supplementary Table S6**). These include two individuals with the AAAAG expansion in *SHANK1* that targeted genotyping delineated to have originated from the paternal side of the family (**Supplementary Figure S2**). These findings further substantiate the contribution of rare TREs to the heritable risk of schizophrenia.

In conclusion, we demonstrate that rare TREs, in particular those that are intronic and close to exons, are an important class of variants contributing to the etiology of schizophrenia. One such example is the TRE in *SHANK1*, a postnatal brain-expressed gene that encodes scaffold proteins that are required for the development and function of neuronal synapses^17^. Genetic variants, including rare CNVs encompassing *SHANK1*, have been observed in individuals with non-syndromic ASD^18,19^ (**Supplementary Information**). Further studies are warranted to delineate the mechanisms of TREs in regulating the expression of *SHANK1* and other TRE-associated genes during brain development. Involvement of genome-wide TREs in schizophrenia may help explain the clinical genetic anticipation that has long been recognized in schizophrenia and suspected to be related to tandem repeats^20^. The enrichment of common variant signals for schizophrenia GWAS at our TRE-associated genes further supports their involvements in schizophrenia. Our findings suggest rare TRE as a potential source of some of the missing heritability for schizophrenia, and highlight the necessity of further genome sequencing studies of TREs in other complex disorders for which missing heritability remains to be identified^21^.

## Data Availability

The 1000G genome-sequencing data are publicly available via Amazon Web Services (s3://1000genomes/1000G_2504_high_coverage/data). Other data produced in the present work are contained in the manuscript.

## Data availability

The 1000G genome-sequencing data are publicly available via Amazon Web Services (s3://1000genomes/1000G_2504_high_coverage/data).

## Code availability

Code used in this manuscript is available from the corresponding author upon reasonable request.

## Acknowledgements

We are grateful to all patients and their families for their participation in this study. We thank The Centre for Applied Genomics (TCAG, a node of CGEn), which is supported by the Canada Foundation of Innovation, Genome Canada, the Hospital for Sick Children, and partners. R.K.C.Y. is supported by The Hospital for Sick Children’s Research Institute, SickKids Catalyst Scholar in Genetics, Brain Canada, The Azrieli Foundation, the University of Toronto McLaughlin Centre and the Nancy E.T. Fahrner Award. This work was also supported by the Canadian Institutes of Health Research (CIHR) (MOP-89066 to A.S.B., MOP-111238 to A.S.B., PJT-175329 to R.K.C.Y.), and a Canada Research Chair in Schizophrenia Genetics and Genomic Disorders (Tier 1, 2009-2016 to A.S.B.). A.S.B. holds the Dalglish Chair in 22q11.2 Deletion Syndrome at the University Health Network and University of Toronto. B. Trost was funded by the Canadian Institutes for Health Research Banting Postdoctoral Fellowship and the Brain Canada Canadian Open Neuroscience Platform Research Scholar Award. B.A.M. was supported by the Restracomp Award from The Hospital for Sick Children

## Contributions

R.K.C.Y. conceived and coordinated the study. W.E. and B.Trost processed genome data for tandem repeat detection and analysis. I.B., L.P., B.H., K.G. and M.K. performed laboratory experiments. W.E., Y.Y. and G.P. performed statistical analyses with additional contributions from B.Trost, and B.Thiruvahindrapuram. Y.Y. and T.H. performed genotype-phenotype correlation and analyses. B.A.M., W.E. and G.P. performed other miscellaneous analyses. A.S.B. managed, recruited, diagnosed and with G.C. examined the recruited participants. R.K.C.Y., B.A.M. and A.S.B. wrote the manuscript with contributions from W.E, B.Trost, I.B., S.W.S., C.R.M., G.C. and C.E.P.. All authors read, reviewed and approved the final manuscript.

## Notes

### Competing Interest Statement

The authors have declared no competing interest.

### Author Declarations

This study was approved by the Research Ethics Board at the Centre for Addiction and Mental Health (CAMH) (151/2002-02) and other local REBs. Written informed consent was obtained for all participants

