## Supplementary information for "Genome-wide tandem repeat expansions contribute to schizophrenia risk"

[1]Genetics and Genome Biology, The Hospital for Sick Children, Toronto, Ontario, Canada
[2]The Centre for Applied Genomics, The Hospital for Sick Children, Toronto, Ontario, Canada
[3]Department of Molecular Genetics, University of Toronto, Toronto, Ontario, Canada
[4]Clinical Genetics Research Program, Centre for Addiction and Mental Health, Toronto, Ontario, Canada
[5]The Dalglish Family 22q Clinic for Adults with 22q11.2 Deletion Syndrome, Toronto General Hospital, University Health Network, Toronto, Ontario, Canada
[6]Division of Clinical and Metabolic Genetics, The Hospital for Sick Children, Toronto, Ontario, Canada
[7]McLaughlin Centre, University of Toronto, Toronto, Ontario, Canada
[8]Genome Diagnostics, Department of Paediatric Laboratory Medicine, The Hospital for Sick Children, Department of Laboratory Medicine and Pathobiology, University of Toronto, Toronto, Ontario, Canada
[9]Department of Psychiatry, University of Toronto, Toronto General Hospital Research Institute and Campbell Family Mental Health Research Institute, Toronto, Ontario, Canada
[*]These authors contributed equally

### Supplementary information

**Clinically relevant rare variants in individuals with schizophrenia**:
Clinically relevant rare variants were defined as small nucleotide changes (single nucleotide variants, and small insertions/deletions) and copy number variants (CNVs) classified as pathogenic/likely pathogenic as potentially clinically relevant and contributing to the expression of schizophrenia, following the guidelines provided by the American College of Medical Genetics and Genomics (ACMG)[1,2] . For complete information on the clinically relevant rare variants please refer to Mojarad et al[3].

**(CTG)n expansions in 3'UTR of *DMPK* and schizophrenia:**
We have previously reported CTG expansions in myotonic dystrophy (DM1)-linked *DMPK* in individuals with schizophrenia and with autism spectrum disorder (ASD)[3,4]. Interestingly, individuals with DM1 had also previously been reported to show signs of schizophrenia, cognitive impairement, social and personality disorders[5-8]. Transgenic mice with >1000 CTG units in *DMPK* show RNA toxicity in the brain, and exhibit neurochemical and electrophysiological signs of synaptic dysfunction, as well as behavioral and cognitive phenotypes[9].

### Supplementary Figures

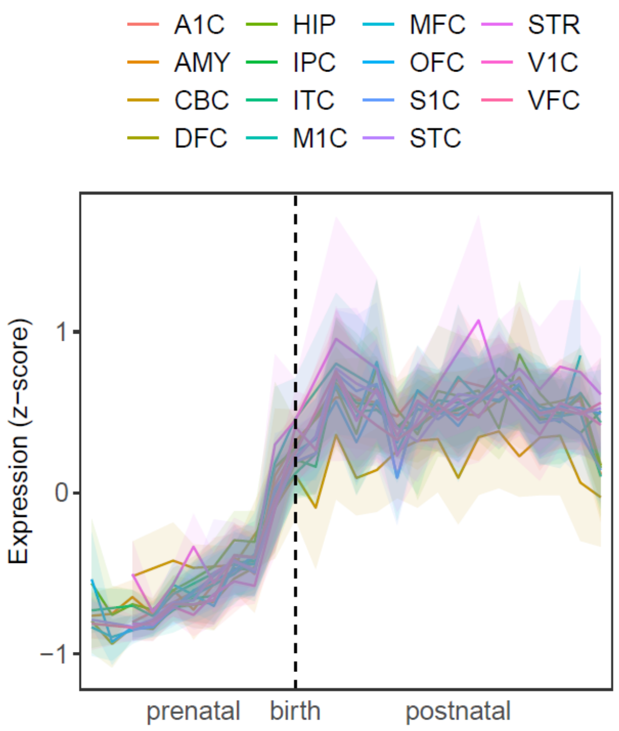


**Supplementary Figure S1. Temporal expression of genes with intronic TREs in different regions of human brain.** Expression in different regions of human brain throughout developmental stages of the postnatally expressed genes (a significantly enriched gene-set, Supplementary Table S4) with intronic TREs in our schizophrenia cohort (Supplementary Table S3). Gene expression for each gene with available data was obtained from the BrainSpan Allen Brain Atlas (http://www.brain-map.org) of Developing Human Brain. Each graph line represents the average expression of 35 genes in each of 15 different brain regions and respective shaded areas represent the 95% confidence intervals of the expression. The results show the intronic TRE gene expression pattern across multiple brain regions.

A1C: primary auditory cortex (core); AMY: amygdaloid complex; CBC: cerebellar cortex; DFC: dorsolateral prefrontal cortex; HIP: hippocampus (hippocampal formation); IPC: posteroventral (inferior) parietal cortex; ITC: inferolateral temporal cortex (area TEv, area 20); M1C: primary motor cortex (area M1, area 4); MFC: anterior (rostral) cingulate (medial prefrontal) cortex; OFC: orbital frontal cortex; S1C: primary somatosensory cortex (area S1, areas 3,1,2); STC: posterior (caudal) superior temporal cortex (area 22c); STR: striatum; V1C: primary visual cortex (striate cortex, area V1/17); VFC: ventrolateral prefrontal cortex.

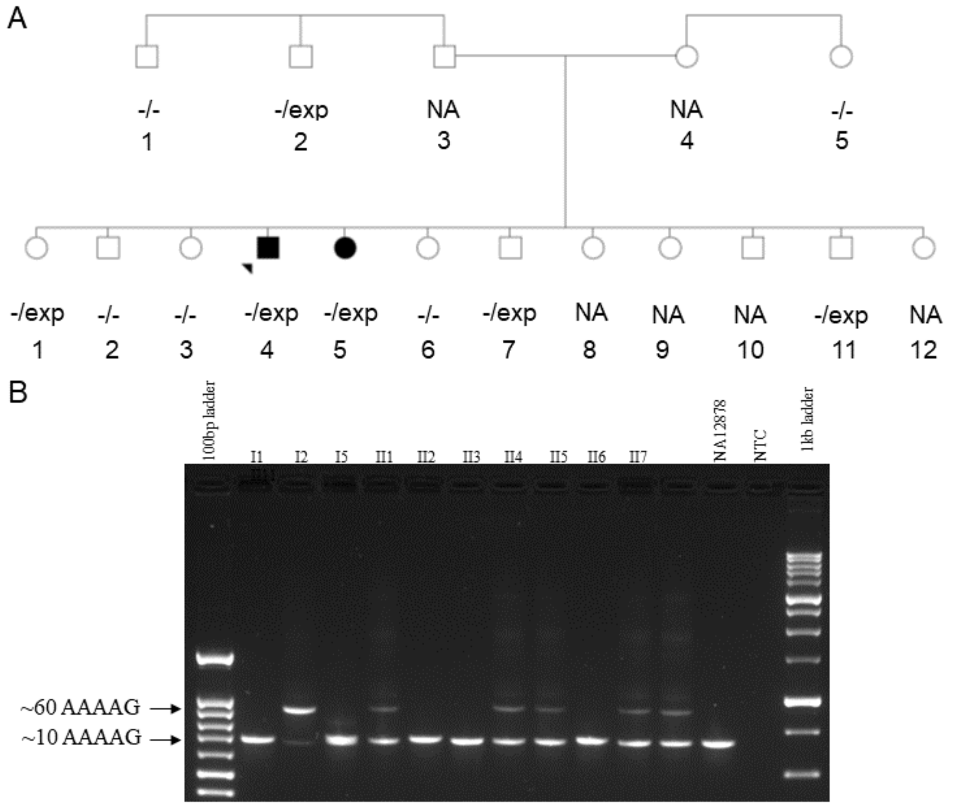


**Supplementary Figure S2. An intronic tandem DNA repeat in *SHANK1* is expanded in an extended family with schizophrenia.** (**A**) Pedigree of family 2 with an (AAAAG)n tandem repeat expansion detected in an intron of *SHANK1* (also identified in an unrelated proband with schizophrenia from the main cohort, Supplementary Tables S1 and S3). Expansions were originally detected using ExpansionHunter Denovo on II4 (proband) and II11 (unaffected sibling), whose DNA was genome-sequenced (Supplementary Table S6). Presence of the *SHANK1* intronic expansion (exp) was then assessed by targeted PCR assay for nine other individuals in the family (Family 2). (**B**) The gel electrophoresis showing one amplicon size (~300bp/~60 additional repeats) larger than those detected in the control DNA (NA12878) corresponding to the intronic TREs identified in *SHANK1* in four of the nine individuals (including II-5 with schizophrenia) from Family 2; DNA was extracted from peripheral blood or lymphoblast cell lines. NA: not available.

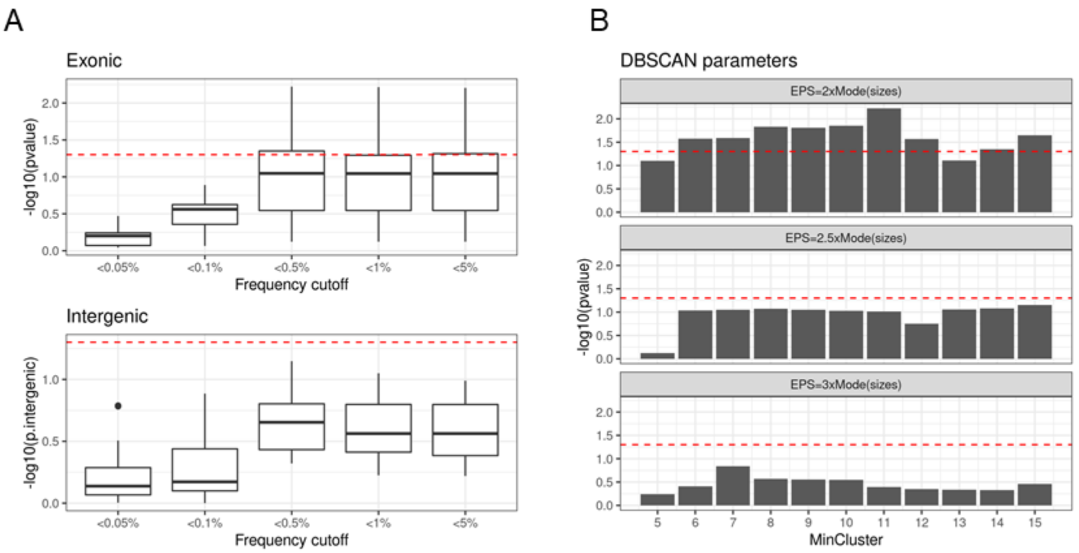


**Supplementary Figure S3. DBSCAN parameter and population frequency cut-off optimization.**
(**A**) Burden test of exonic (i.e., exon-proximal) and intergenic TREs at different population frequency cut-offs. Data points in box plots were based on different sets of rare TREs identified using 33 different DBSCAN parameter combinations (Methods). (**B**) DBSCAN minimum cluster member and Epsilon parameters optimization using <0.5% as the population frequency cut-off. Bars indicate p-value of the test of rare exonic tandem repeats burden. Dashed red line represents p=0.05. Raw data are provided in Supplemental Table S8.

### Online Methods

#### Ethics Statement

This study was approved by the Research Ethics Board at the Centre for Addiction and Mental Health (CAMH) (151/2002-02) and other local REBs. Written informed consent was obtained for all participants[3].

#### Samples sequencing, and genome alignment

We used genome sequencing data from Canadian individuals of European descent (257 with schizophrenia, and 225 with congenital heart disease (CHD) and no psychotic illness), as well as 2,504 samples from the 1000 Genomes Project (1000G)[10]. The schizophrenia samples and non-psychiatric controls were assessed for quality, and prepared using TruSeq DNA library prep kits. These samples were sequenced on the Illumina HiSeq X platform (2×150-bp paired-end reads) at The Centre for Applied Genomics (TCAG, Toronto, Canada) and processed for alignment and genomic variant calling as previously described[3,11]. The 1000G samples were sequenced on the Illumina NovaSeq platform (2×150-bp paired-end reads). The 1000G genome sequencing data are publicly available and we downloaded them via Amazon Web Services (s3://1000genomes/1000G_2504_high_coverage/data). All samples were aligned to the GRCh38/hg38 reference genome using BWA-mem[12]. The study protocol was approved by the

Research Ethics Boards of The Hospital for Sick Children and Centre for Addiction and Mental Health. Informed consent was obtained from all participants at the recruitment locations.

**Genome-wide identification of tandem repeats**

Genome-wide detection of tandem repeats was performed as previously described[4]. Briefly, we used ExpansionHunter Denovo (EHdn; https://github.com/Illumina/ExpansionHunterDenovo)[13] to estimate the size and location of genomic tandem repeats. For a tandem repeat to be detected by EHdn, it must be larger than the sequence read length (for example, >150 bp). As a result, samples that did not meet this minimum size for a given region were left without size estimation by EHdn. We compared the TRs identified by EHdn to tandem repeats in the human reference genome, derived from Tandem Repeats Finder (TRF)[14]. To support the accuracy of EHdn-predicted tandem repeat sizes, we genotyped the 8 rare exon-proximal repeat loci (9 motifs) identified in 13 individuals, along with 6 selected rare intronic repeat loci, using ExpansionHunter v.3.0.2[15,16], which estimates allele-specific tandem repeat sizes for each genomic coordinate and motif supplied by the user with high accuracy (precision = 0.91, recall = 0.99)[15,16]. To determine more precise coordinates for input to ExpansionHunter, we identified coordinates from TRF that overlapped each locus. For each combination of TRF coordinates and EHdn motif, we used ExpansionHunter to estimate motif-specific (as detected by EHdn) tandem repeat sizes for the samples involved. We then calculated the Spearman correlation coefficient and P value between the EHdn-predicted tandem repeat sizes and the size estimated by ExpansionHunter (defined as either the size of the longest allele or the sum of the two allele sizes), aggregated over all of the EHdn-detected motifs for that locus (**Supplementary Table S7**). We also manually evaluated the presence of tandem repeat expansions and the corresponding motif by inspecting reads from the BAM file for tandem repeats that were found to be expanded by EHdn for all.

**Detection of rare tandem repeat expansions and sample quality assessment**

We excluded tandem repeats with different size distribution between schizophrenia and CHD samples (two-sided Wilcoxon's signed-rank test $P < 0.05$) in order to avoid any potential technical biases in estimating tandem repeat size between the two cohorts. To detect rare tandem repeat expansions, we followed Trost et al.[4], by applying the non-parametric Density-Based Spatial Clustering of Applications with Noise (DBSCAN) algorithm to identify outliers based on EHdn estimated tandem repeat size in each locus. We optimized two DBSCAN parameters as well as the population frequency cut-off for rare TRE identification (**Supplementary Figure S3**; **Supplementary Table S8**). Based on a different set of rare TREs identified using different DBSCAN parameters and population frequency cut-off, burden test of exonic TREs and intergenic TREs were performed with the total number of rare TREs as a covariate. The DBSCAN parameters set (minclust = 11, eps = 2 x mode of EHdn sizes) and population frequency cut-off (frequency < 0.05) that provided the strongest signal in the exonic TREs burden test and weakest signal in intergenic TREs burden test was selected for the next step of the analysis. Sample quality assessment was done by inspecting the counts of total number of tandem repeat loci and TREs per sample. Anscombe transformation was done on the counts to put the count distribution closer to the gaussian distribution. Three control samples and five schizophrenia samples with the transformed counts exceeding 3 standard deviations from the mean of the transformed counts were tagged as outliers and excluded from the analysis. Rare TRE identification was then performed on the remaining set.

**Burden analysis**

To compare the prevalence of rare TREs in individuals with and without schizophrenia, we performed a logistic regression analysis by regressing the number of rare tandem repeat expansions on the affected status (unaffected = 0, affected = 1). For this analysis, we only included tandem repeats on autosomal chromosomes to avoid sex bias. Biological sex and the total number of genomic TRs per individual were used as covariates. To test the burden of TREs in different functional elements (for example, exons and introns), we separated the genome (RefSeq, GRCh38) into different functional elements: upstream (1 kb upstream of the transcriptional start site, TSS), 5′ untranslated regions (5' UTR), exon, core splice site, intron, 3′ UTR and downstream (1 kb downstream of transcription termination sites)[4]. If any rare TRE affected more than one functional element, we prioritized the effects based on their impact on the corresponding genes predicted by ANNOVAR (October 2019 release)[17].

**Experimental validation of tandem repeat expansions in *SHANK1***

Validation of the *SHANK1* TREs detected by EHdn was completed by PCR, gel electrophoresis, and Sanger sequencing. We designed primers flanking the repeat of interest with the following sequences: 5'-CCTATCTCCTATGAATGGACGAC-3' and 5'-GATGCCGTTAAATGCGAGTTTC-3'. We performed PCR on the samples using HotStarTaq DNA polymerase (Qiagen), a primer annealing temperature of 63°C, and an elongation time of 2 minutes. We then ran the PCR products on a 1.2% agarose gel to confirm the size of the repeats, and performed Sanger sequencing to confirm the sequence. Other DNA samples from this cohort without a predicted expansion in *SHANK1* were run under the same conditions as negative controls (Supplementary Figure S2).

**Statistical comparison of clinical features**

We hypothesized that individuals with schizophrenia compared with CHD and no psychotic illness (**Figure 1A and B**), and differentially within schizophrenia those with clinical features, including family history of schizophrenia in first degree relatives, intellectual disability and syndromic features (**Figure 2A and 2C**), would have a greater contribution from the genic and exonic (i.e., exon-proximal, within 300 bp of exon junctions) TREs identified. Therefore, we used the non-parametric one-sided Wilcoxon signed-rank test to compare the datasets unless stated otherwise.

**Enrichment in common variant risk**

For the 193 TRE-associated genes that we identified in this schizophrenia sample, we used MAGMA v.1.09b11 as described previously[18] to determine whether they were enriched in common variant risk loci for schizophrenia and other traits. Specifically, we compared our 193 TRE-associated gene set against summary statistics from genome-wide association studies for schizophrenia[19], ASD[20], attention deficit/hyperactivity disorder[21], educational attainment[22] and (as a negative control) height[23] (**Figure 2B**, **Supplementary Table S9**). We also tested for an enrichment of 193 TRE-associated genes in the 655 genes within 270 refined genome-wide significant loci that involve fewer than 4 causal variants in the latest schizophrenia GWAS of 69,369 cases and 236,642 controls[24]. We applied one-sided Fisher's Exact test to compare the enrichment of GWAS signals in TRE-associated genes (n=193) against other genes that are not associated with rare TRE (n=19,276).
